# Altered visual evoked potentials associated with verbal and nonverbal skills in Fragile X syndrome

**DOI:** 10.1101/2022.07.10.22277277

**Authors:** Elizabeth Saoud, John Fitzgerald, Megan Hartney, Carol L. Wilkinson

**Author notes:** To whom correspondence should be addressed: Carol Wilkinson, Labs of Cognitive Neuroscience, 2 Brookline Place, Brookline, MA 02445.

## Abstract

**Background:** Understanding the neurobiology behind Fragile X syndrome (FXS) is critical in identifying effective therapeutics and improving care for affected individuals.

Electroencephalography (EEG) provides an opportunity to investigate the biological foundations of this disorder. We aimed to characterize the visual evoked potential (VEP) in young children with FXS, and to understand how measures of the VEP are associated with verbal and nonverbal development within FXS.

**Methods:** VEPs were collected in children between 2-7 years old with FXS (n = 9) as well as corresponding age-(n = 10) and cognitive-matched (n = 9) typically developing children. Additionally, the Mullen Scales of Early Learning and Preschool Language Scales were administered to collect measures of verbal and nonverbal development. Differences in component amplitudes and latencies of the VEP were assessed using ANCOVAs, and associations of VEP measures and verbal and nonverbal development were evaluated using linear regression with age as a covariate.

**Results:** No differences between groups were observed in N1, P1, or N2 VEP components. However, a consistent and prominent P2 component (latency = 177ms ± 13.7), was observed in children with FXS. The P2 amplitude was significantly increased in FXS children compared to the cognitive-matched group (p = 0.004). For children with FXS, the amplitude of several VEP components were associated with verbal and nonverbal development; larger N1 amplitude and smaller P1 and P2 amplitudes were all associated with better receptive language (all p<0.05) and larger N1 amplitude was also associated with better fine motor skills (p<0.05).

**Conclusions:** The observed increase in P2 amplitude and its negative association with language development within the FXS group supports the P2 component as a potential biomarker for FXS as a disorder, as well as a pathophysiological marker of verbal impairment that could be used in clinical trials.

## Background

Fragile X Syndrome (FXS) is the most common inherited form of intellectual disability, and children with FXS often struggle with severe language impairments as well as sensory and behavioral challenges (Hagerman et al., 2017). FXS is caused by a CGG trinucleotide repeat expansion on the X chromosome, leading to silencing of the *Fmr1* gene. This reduces production of Fragile X Messenger Ribonucleoprotein 1 (FMRP), leading to overexpression of other local proteins that negatively impact synapse development, strength, and plasticity (Garber et al., 2008). The cognitive and behavioral deficits experienced by children with FXS overlap with other neurodevelopmental disorders. For example, around 50% of males with FXS meet criteria for autism spectrum disorder (CDC, 2021), and many if not the majority of children with FXS meet criteria for attention deficit hyperactivity disorder (ADHD) and anxiety (Hagerman et al., 2009). The neuropathology underlying FXS points to an imbalance of neural excitation and inhibition (Contractor et al., 2015), which likely disrupts downstream synaptic and homeostatic plasticity and ultimately learning and memory. Mouse models of FXS exhibit reduced GABA transmission, resulting in over-excitation (Gibson et al., 2008; Lozano et al., 2014), and several GABA agonists have successfully improved physiological and behavioral phenotypes in mice (Henderson et al., 2012; Qin et al., 2015). Unfortunately, most human phase II trials have not been successful in part due to the lack of validated brain-based biomarkers to monitor clinical improvement (Berry-Kravis et al., 2013; Jacquemont et al., 2014).

The visual evoked potential (VEP) has potential as a brain-based biomarker candidate in neurodevelopmental disorders as it has been used in both human and mouse studies, and is non-invasive, low cost, and feasibly collected across the lifespan from infants to adults. As a neural measure, the VEP provides information both about integrity of the visual pathway and higher-level cognitive processing of sensory information. As such, the VEP can be used as a neurological biomarker, inform underlying neurophysiology, and map progression of disorders that affect sensory systems (Odom et al., 2004). Several studies have examined differences in VEP across several neurodevelopmental disorders, including FXS, Down syndrome, 16p11.2 variants, tuberous sclerosis complex (TSC), and Rett syndrome. Alterations in component amplitudes have been observed across disorders, with increased P1 amplitude in 16p11.2 deletions (LeBlanc & Nelson, 2016) and TSC (Varcin et al., 2016), and decreased P1 amplitude in 16p11.2 duplications (LeBlanc & Nelson, 2016) and Rett syndrome (LeBlanc et al., 2015). In FXS, a study of VEP response in individuals 10-22 years old observed increased N1 and N2 amplitude when compared to age-matched, but not cognitive matched comparison groups (Knoth et al., 2014). Despite differences being observed in a range of neurodevelopmental disorders, few studies have examined the relationship between VEP responses and behavioral and cognitive outcomes – a crucial component of biomarker identification. Further in FXS, there have been few EEG studies in young children; identification of EEG based biomarkers associated with developmental outcomes at these young ages is crucial to biomarker efficacy in clinical trials for young children.

This study had two main aims. First, we characterize the VEP in children with FXS compared to both age-and cognitive-matched comparison groups. We hypothesized that FXS children would have differences in their VEP components compared to both groups. Second, within the FXS group we assess how VEP measures are associated with varying levels of cognitive and language development.

## Methods

### Participants

Table 1 describes participant characteristics. A total of 16 FXS children (age 33-78 months old) with full mutation of *Fmr1* and 12 similarly aged typically developing children were recruited for this study (IRB#P00025493) conducted at the Labs of Cognitive Neuroscience at Boston Children’s Hospital. 6 FXS participants and 1 age-matched participant did not complete VEP EEG acquisition. VEP data from 1 FXS and 1 age-matched participant was excluded due to excessive artifact. VEP and behavioral data was analyzed for a total of 9 FXS participants and 10 age-control participants.

**Table 1:** Demographics of Subjects

|  | Age-Matched<br>(N = 10) | FXS<br>(N = 9) | Cognitive-Matched<br>(N = 9) |
| --- | --- | --- | --- |
| **Age, mean in months (SD)** | 48.6 (14.1) | 44.3 (14.5) | 25.2 (8.5) |
| **Highest parental education, n (%)** |  |  |  |
| ≤ High school/GED | 0 (0) | 0 (0) | 0 (0) |
| Associate's degree | 0 (0) | 1 (11) | 0 (0) |
| Bachelor's degree | 1 (10) | 2 (22) | 1 (11) |
| Graduate or professional degree(s) | 9 (90) | 6 (67) | 8 (89) |
| **Household income, n (%)** |  |  |  |
| < \$40,000 | 0 (0) | 0 (0) | 0 (0) |
| \$40-70,000 | 0 (0) | 3 (33) | 1 (11) |
| >\$70,000 | 10 (100) | 6 (67) | 8 (89) |
| No response | 0 (0) | 0 (0) | 0 (0) |
| **Sex, n (%)** |  |  |  |
| Female | 0 (0) | 1 (11) | 3 (33) |
| Male | 10 (100) | 8 (89) | 6 (67) |
| **Race, n (%)** |  |  |  |
| White | 6 (60) | 6 (67) | 6 (67) |
| Black/African American | 0 (0) | 0 (0) | 0 (0) |
| Asian | 0 (0) | 1 (11) | 0 (0) |
| Multi-race | 4 (40) | 1 (11) | 3 (33) |
| No response | 0 (0) | 1 (11) | 0 (0) |
| **Ethnicity, n (%)** |  |  |  |
| Hispanic or Latino | 0 (0) | 2 (22) | 2 (22) |
| **Mullen Scales of Early Learning (mean ± SD)** |  |  |  |
| Nonverbal Developmental Quotient | 109.1 ± 11.0 | 59.1 ± 13.6 | 110.1 ± 17.8 |
| Visual Reception Raw Score | 44.8 ± 3.4 | 28.4 ± 7.2 | 28.0 ± 5.5 |
| Fine Motor Raw Score | 38.0 ± 4.5 | 24.4 ± 4.2 | 25.6 ± 5.3 |
| **EEG Quality Measures (mean ± SD)** |  |  |  |
| Number of Trials Post Rejection | 91.3 ± 9.8 | 76.4 ± 20.1 | 91.2 ± 11.1 |
| Percent Good Channels | 80.6 ± 10.8 | 80.9 ± 6.1 | 87.7 ± 7.8 |
| 1 Hz Pre and Post Wav-Thresholding Correlation | 0.72 ± 0.17 | 0.66 ± 0.15 | 0.86 ± 0.05 |

#### FXS and age-controls

FXS participants all had documented full mutation of the *Fmr1* gene; however, methylation status was not known for all participants. All data was collected prior to the COVID pandemic, which halted additional recruitment. Prior to this, 1 female participant with FXS had been enrolled, but we had not yet enrolled an age-and sex-matched comparison participant. Therefore, the age-matched comparison groups were composed of all males. Across FXS and age-matched groups, additional exclusion criteria included history of prematurity (< 35 weeks gestational age), low birth weight (< 2000 g), known birth trauma, known genetic disorders, unstable seizure disorder, current use of anticonvulsant medication, and uncorrected hearing or vision problems. Children were required to be from a primarily English-speaking household with English spoken more than 50% of the time.

#### Cognitive matched controls

VEP data from an additional 9 cognitive-matched children were analyzed. Individuals from this group provided data as part of a concurrent longitudinal study (IRB#P00018377) in infants 12-36 months old which used the same VEP paradigm in the same EEG rooms. Exclusion criteria were the same as above. Participants were matched based on raw Fine Motor and Visual Reception scores on the Mullen Scales of Early Learning (see Methods: Developmental Assessment).

Institutional review board approval was obtained prior to starting the study, and written, informed consent was obtained from all parents or guardians prior to their child’s participation.

### Developmental Assessment

Nonverbal subscales (Fine Motor and Visual Reception) of the Mullen Scales of Early Learning (MSEL; Mullen, 1995) were administered to all FXS and cognitive-matched participants, and age-matched controls under 70 months of age. The Preschool Language Scale 5th Edition (PLS; Zimmerman et al., 2011) was administered to FXS and age-matched controls. The PLS assesses early language development, including both receptive and expressive language skills. We utilized the raw scores from the Expressive Communication and Auditory Comprehension subscales.

### VEP collection

EEG data were collected in a dimly lit, sound-attenuated, electrically shielded room. The child sat either in their caregiver’s lap or independently in a chair or stroller while looking at a Tobii 120 monitor (Tobii Technology, Sweden) approximately 60cm away, and caregivers were instructed to avoid social interactions with their child. A research assistant was also present in the room to help keep the child calm and focused, if necessary. Pattern-reversal visual stimuli, consisting of a reversing black and white checkerboard pattern (with diagonal check size of 0.5cpd), were presented on the monitor using ePrime software (Psychology Software Tools Inc., Pittsburgh, PA, USA). Stimuli were presented for at least 500ms as the stimulus phase-reversal was contingent on binocular fixation on the monitor for a period of 100ms to ensure the participant was looking for each trial. In a small number of cases, the eye tracking software did not accurately pick up on the child’s binocular fixation. In these cases, the research assistant manual progressed the stimuli based on whether the child was looking at the screen. A maximum of 200 trials were presented, with fewer trials presented if the child became inattentive. Continuous EEG was recorded using a 128-channel high density HydroCel Geodesic Sensor Net and amplified with a NetAmps 300 high-input amplifier (Electrical Geodesics Inc., Eugene, OR, USA). Data were sampled at 1000Hz, and impedance was kept below 100kOhms in accordance with the impedance capabilities of the high-impedance amplifiers inside the electrically shielded room (Ferree et al., 2001).

### EEG pre-processing

Raw Netstation files were pre-processed, and artifact was removed using the Harvard Automated Processing Pipeline for EEG plus Event-Related Software (HAPPE+ER; Monachino et al., 2021). Line noise was removed using Cleanline’s multi-taper regression, data was low pass filtered at 100Hz, and bad channels were removed. Artifact was then extracted using wavelet-thresholding with wavelet resolution level set to ≤0.1Hz as determined to be optimal in infant VEP analyses by Monachino et al. 2021. Data was then filtered (0.3-30Hz) and segmented (−100 to 400ms) around the visual stimulus, and baseline corrected via mean subtraction. Segments with retained artifact in the region of interest used for VEP analysis (Electrodes: O1, O2, 71, 75, and 76) were then rejected using HAPPE+ER’s amplitude and joint probability method with an artifact threshold of ±200 μV. Bad channels were then interpolated and data was referenced to the average reference.

### EEG rejection criteria

EEGs were excluded from the final sample if (1) they had fewer than 20 trials post segment rejection, (2) >3/5 electrodes in the region of interest were interpolated, or (3) they did not meet the following HAPPE+ER quality metrics: cross-correlation value < 0.2 for pre/post wavelet thresholding for frequencies 0.5, 1, and 2Hz. 1 FXS and 1 age-matched participant were excluded based on the above criteria.

### VEP analysis

Grand average waveforms were calculated by averaging the 5 occipital electrodes described above. Given observed differences in latency of components between groups, components were identified as follows: The P1 component was identified as the most positive peak between 80-120ms after stimulus onset. The N1 and N2 components were then identified as the relative minimums that come before and after the P1, respectively. The P2 component was identified as the relative maximum after the N2. Finally, N1-P1 amplitude was calculated as the amplitude of the P1 measured from the preceding N1 peak, and P1-N2 amplitude was calculated as the amplitude of the N2 measured from the preceding P1 peak. Individual VEPs were visually inspected to confirm or adjust VEP components.

### Statistical analysis

ANCOVAs with post-hoc pairwise Tukey HDR tests were conducted to determine if there were significant differences in the mean VEP amplitude and latencies between groups, while accounting for covariates related to data quality, specifically number of trials post-rejection and 1Hz pre-and post-thresholding correlation. A false discovery rate (FDR) correction was used to account for the multiple comparisons made for each analysis.

Linear regression was used to determine if component amplitudes were associated with nonverbal and language measures within the FXS group. Raw scores were used, and age was included as a covariate, as both VEP and raw scores on behavioral tests change with age. Analyses were performed in Python 3.9 with Statsmodels and Pingouin. Figures were created using python data visualization libraries [matplotlib (Hunter, 2007) and Seaborn (https://seaborn.pydata.org/index.html)]

## Results

### Sample Description

A summary of the demographic information and mean scores for MSEL nonverbal scales are shown in Table 1. As expected, the mean nonverbal developmental quotient (NVDQ) was lower for the FXS group, reflecting developmental delays associated with this syndrome. Raw scores on subscales were well matched between FXS and cognitive-matched controls. EEG quality measures are also shown in Table 1. The FXS group had a significantly smaller number of trials post-rejection (ANOVA, p = 0.048) and a significantly smaller 1Hz pre-and post-thresholding correlation (ANOVA, p = 0.004) when compared to its control groups. This latter value suggests that EEG data collected from the FXS group, had greater signal change before and after wavelet thresholding likely due to increased noise in the data, although mean values across all measures were well above our exclusion criteria. Given the significant differences between groups, EEG quality measures were used as co-variates in statistical analyses.

### VEP Waveform

Grand-averaged VEPs for the FXS, age-matched, and cognitive-matched groups are shown in Figure 1. All groups demonstrated a typical N1-P1-N2 pattern response. However, a prominent P2 was also observed in the FXS group. Statistical analyses (Table 2) confirmed a significant group effect for P2 amplitude, and post-hoc pairwise Tukey HSD tests demonstrated FXS children having significantly increased P2 amplitude compared to the cognitive-matched (p = 0.004, Figure 1) comparison group. A similar trend was observed when compared to the age-matched comparison group, but with marginal significance (p = 0.052). While there was no significant difference in P2 latency, we observed more consistent timing of the P2 within the FXS group (Coefficient of variance FXS = 0.07, Age-Matched = 0.12; Cog-Matched = 0.14; Figure 1). No other components showed significant differences between groups.

**Table 2:** Summary of Amplitude and Latencies of Components.

|  | Age-Matched,<br>mean ± SD | FXS,<br>mean ± SD | Cognitive-Matched,<br>mean ± SD | ANCOVA,<br>F | P Value,<br>unadjusted;<br>adjusted |
| --- | --- | --- | --- | --- | --- |
| **Component Amplitude (µV)** |  |  |  |  |  |
| **N1 Amplitude** | -4.56 ± 3.48 | -6.77 ± 6.00 | -6.24 ± 4.16 | 1.518 | 0.240; 0.321 |
| **P1 Amplitude** | 3.63 ± 4.29 | 6.71 ± 6.97 | 3.13 ± 4.05 | 1.128 | 0.341; 0.379 |
| **N2 Amplitude** | -2.32 ± 5.04 | 0.80 ± 8.42 | -7.97 ± 6.09 | 2.496 | 0.105; 0.261 |
| **P2 Amplitude** | 0.95 ± 5.39 | 8.63 ± 5.62 | -1.48 ± 5.14 | 6.769 | **0.005; 0.049** |
| **N1P1 Amplitude** | 8.19 ± 5.04 | 13.5 ± 7.26 | 9.37 ± 6.01 | 2.648 | 0.092; 0.261 |
| **P1N2 Amplitude** | 5.95 ± 4.50 | 5.91 ± 3.64 | 11.1 ± 6.81 | 1.630 | 0.218; 0.321 |
| **Latency (ms)** |  |  |  |  |  |
| **N1 Latency** | 56.6 ± 9.29 | 64.2 ± 4.94 | 63.8 ± 9.46 | 2.789 | 0.082; 0.261 |
| **P1 Latency** | 93.4 ± 4.43 | 97.1 ± 8.95 | 96.0 ± 6.08 | 0.445 | 0.646; 0.646 |
| **N2 Latency** | 126 ± 13.6 | 127 ± 11.5 | 138 ± 14.2 | 1.499 | 0.245; 0.321 |
| **P2 Latency** | 160 ± 20.0 | 177 ± 13.7 | 182 ± 27.3 | 1.442 | 0.257; 0.321 |

**Figure 1:**
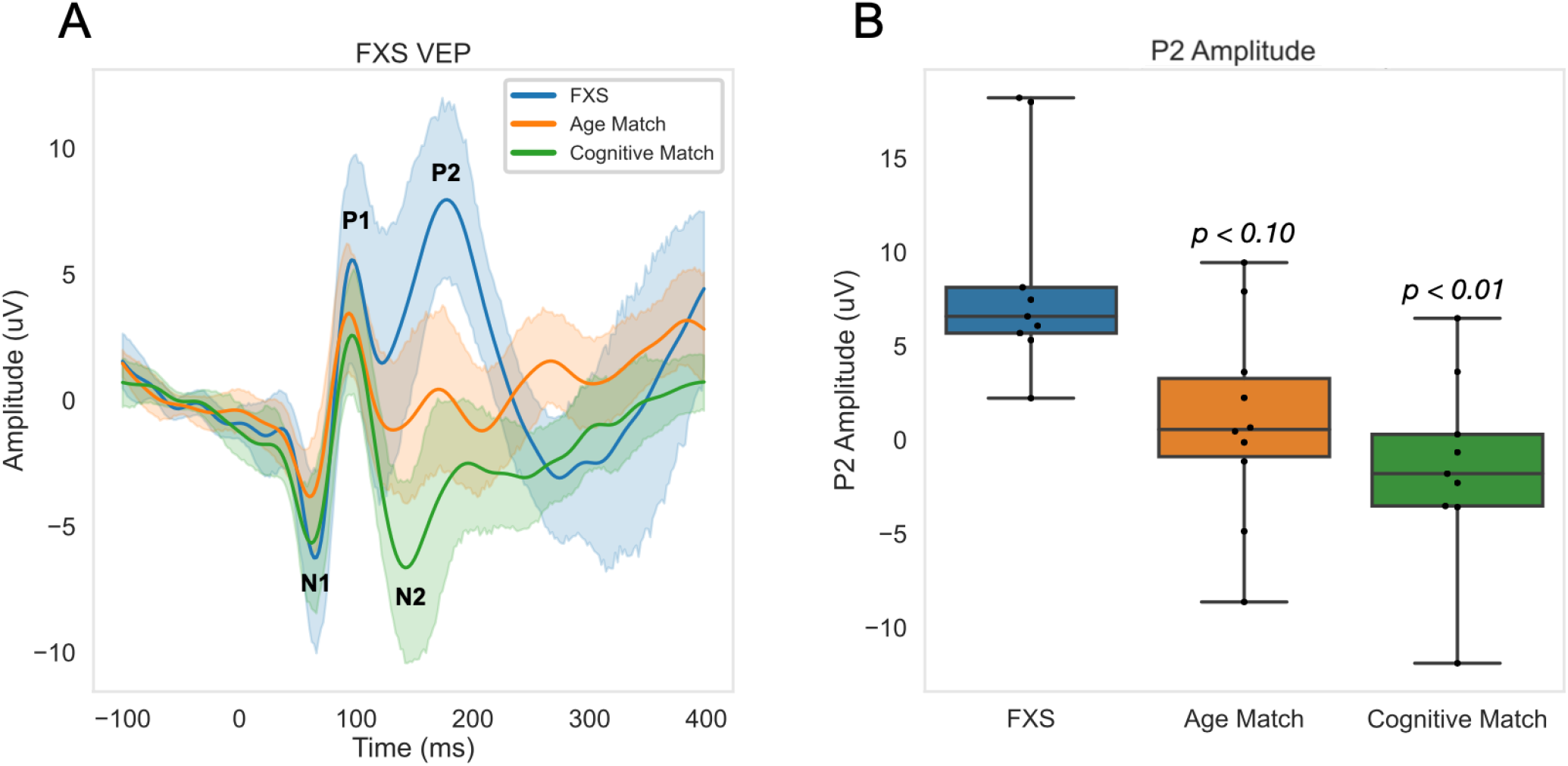
(A) Grand-averaged VEPs are shown, with 95% confidence intervals. (B) Mean P2 Amplitude across trials across groups. P values shown are in comparison to the FXS group.

### VEP and Nonverbal Development

We next examined whether component amplitudes were associated with measures of nonverbal development, specifically the MSEL visual reception and fine motor subscales. Results from linear regression analyses with age included as a covariate are shown in Table 3. Within the FXS group, a larger, more negative N1 was correlated with higher fine motor scores. All other associations were not significant. Visualization of unadjusted Pearson correlation between N1 and fine motor scores is shown in Figure 2.

**Table 3:**
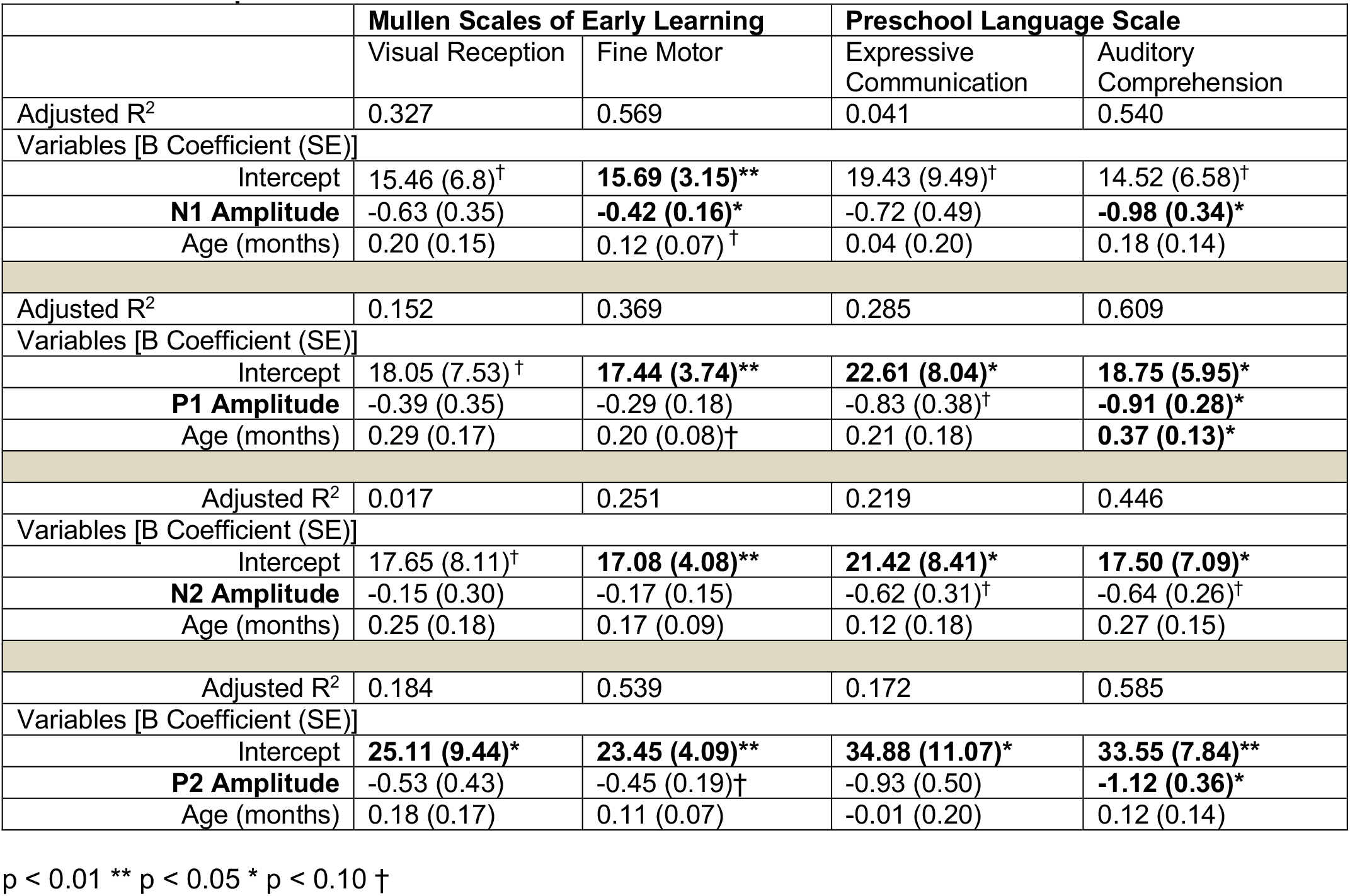
VEP component associations with clinical measures.

**Figure 2:**
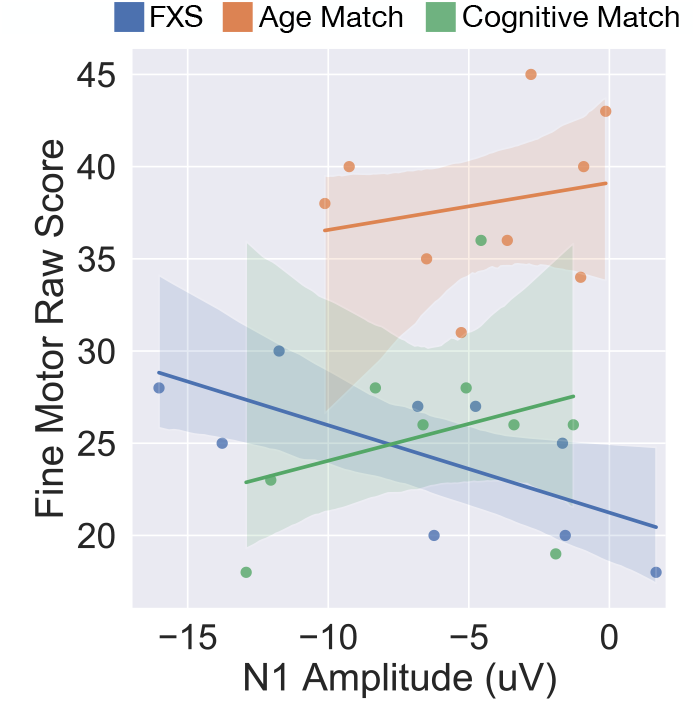
Correlation between N1 Amplitude and MSEL fine motor raw score for FXS, Age Match, and Cognitive Match groups.

### VEP and Verbal Development

We next examined associations between component amplitudes and language development, as measured using the PLS-5. Linear regressions with age as a covariate found that larger N1 amplitude, and smaller P1 and P2 amplitudes were associated with better receptive language skills (p = 0.028, p = 0.017, p = 0.020, respectively; Table 3). Additionally, association between the N2 amplitude and Auditory Comprehension raw score was marginally significant (p = 0.052). Only marginally significant associations were found between component amplitudes and the Expressive Communication subscale. Visualization of unadjusted Pearson correlations are shown in Figure 3. Note that the PLS-5 was only administered to FXS and age-matched controls.

**Figure 3:**
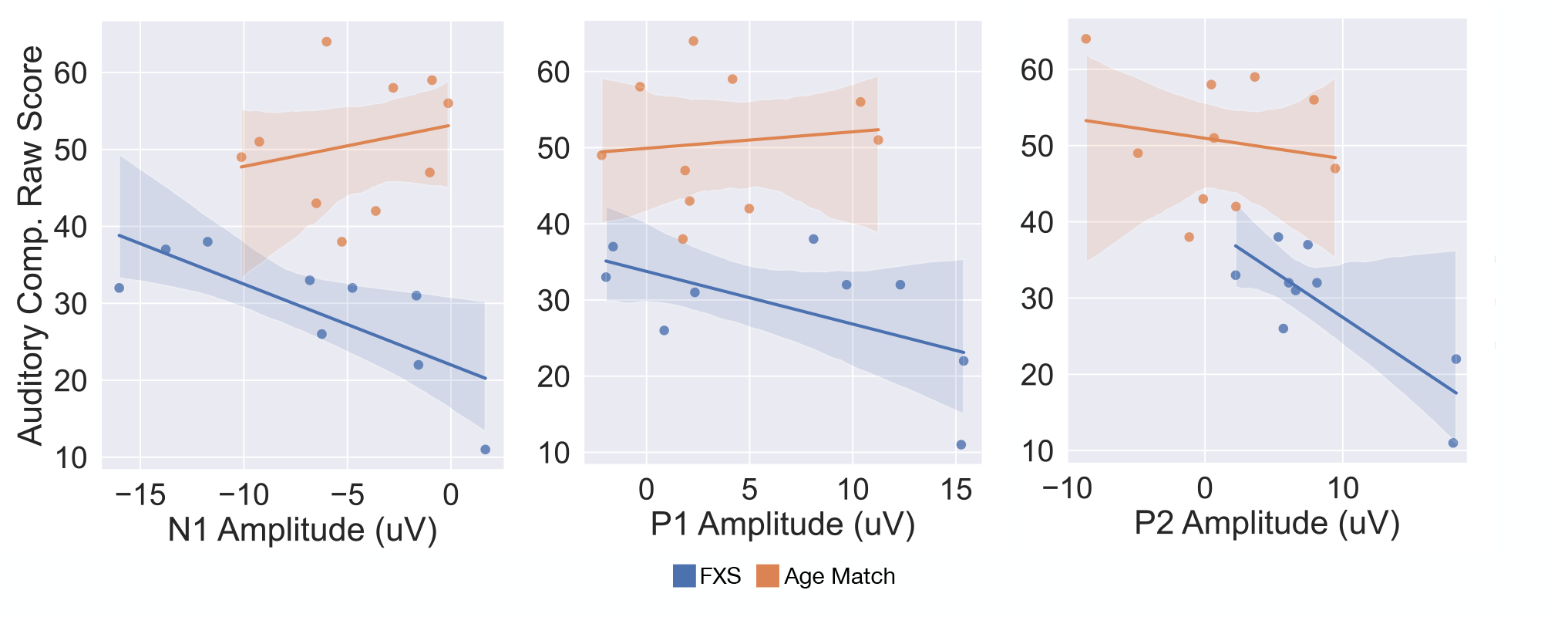
Correlations between N1, P1, and P2 Amplitude and PLS Auditory Comprehension Raw score for FXS and Age Match groups.

## Discussion

In this study, we compared the VEP response in children with FXS compared to both age-and cognitive-matched typically developing children. Overall, we observed that children with FXS showed an increased P2 amplitude which was negatively associated with receptive language development. These findings support further investigation of the P2 component as a potential biomarker for the FXS population.

### VEP Response

The FXS group produced clear and robust early VEP responses that largely matched those of typically developing children, indicating that early-and lower-order visual processing is likely functionally intact for this group. Instead, differences in a later and higher-order visual processing component, the P2, was observed. FXS participants displayed an increased P2 amplitude compared to the cognitive-matched comparison group, with a similar but not-significant trend when compared to the age-matched group. Prior ERP studies in male adults with FXS have observed increased P2 amplitude in response to auditory but not in response to visual stimuli (Knoth & Lippé, 2012; Van der Molen et al., 2012). In another study specifically using a checkboard VEP stimulus, Knoth et al. also did not observe differences in P2 amplitude. However, this study was in an older population (10-22 years) that included a larger percentage of females who, although had full mutation, were also mosaic, which is associated with less severe symptom presentation (Knoth et al., 2014). The VEP P2 component has been less well studied than early components, but is thought to represent both primary sensory and higher cognitive processing. It has been suggested that P2 amplitude is negatively associated with attention. However, as our task was passive, we do not have a simultaneous measure of attention to determine whether decreased attention was associated with increased P2 amplitude in this sample. There is some evidence that P2 response in a visual priming task in adults is associated with theta (4-6Hz) phase-locking (Freunberger et al., 2007), and interestingly, studies in FXS adults have observed increased theta power in response to auditory stimuli compared to age-matched adults without FXS (Ethridge et al., 2019).

Differences in VEP component amplitudes have also been observed in several genetic neurodevelopmental disorders including tuberous sclerosis complex and 16p11.2 copy number variants (LeBlanc & Nelson, 2016; Varcin et al., 2016). Most of these studies identify differences in the early components, namely the N1 and P1, which are hypothesized to be related to early processing of sensory information. In our FXS participants we observed only a marginally significant group effect for the N1-P1 amplitude prior to adjusting for multiple comparisons, with the FXS group having a larger N1-P1 amplitude compared to comparison groups. The P2 component has not been extensively studied in other neurodevelopmental disorders. However, we do note that VEP waveforms obtained in Rett syndrome patients visually appear to have an increased P2 amplitude, although this relationship was not statistically assessed (LeBlanc et al., 2015). Identifying genetic disorders with shared ERP findings could shed further light on the underlying mechanisms of the altered P2 component.

### VEP Associations with Nonverbal and Verbal Measures

Previous studies have examined correlations between VEP component amplitudes and various measures of development, finding that increases in component amplitudes are often associated with better outcomes in typically developing populations (Jensen et al., 2019; Josiassen et al., 1988). We hypothesized that N1 and P1 amplitudes would be associated with nonverbal developmental measures. Indeed, we observed that within the FXS group a larger (more negative) N1 amplitude was associated with better fine motor skills after adjusting for age. Significant associations between other components and non-verbal measures were not observed. In addition, we did not observe significant associations between VEP components and nonverbal development within the two typically developing samples, although our sample size is small, so negative findings must be interpreted with caution.

We also examined associations between component amplitudes and measures of verbal development. Here, a larger (more negative) N1 amplitude and smaller (less positive) P1 and P2 amplitudes were significantly associated with better receptive language within the FXS group. Association between N2 amplitude and receptive language was marginally significant. Associations between VEP measures and expressive communication were not found to be significant, although the trends were consistent across these measures for FXS children. Our finding that both early (N1) and later (P2) components are associated with receptive language, could be interpreted in several ways. First, it may suggest that alterations in visual sensory processing affect language development, with receptive language processing being more affected due to its increased reliance on additional visual information, especially at young ages. Alternatively, these findings may instead reflect global network alterations within FXS, such as imbalances in glutamatergic/GABAergic signaling, separately affecting visual brain responses and language development. In this same cohort of subjects, we found that increased aperiodic gamma power, hypothesized to be a marker increased cortical excitation, was associated with better language abilities (Wilkinson & Nelson, 2021). Differences in basal neuronal network excitability could cause differences in VEP response. Exploratory correlation analysis between baseline aperiodic gamma power VEP components found no significant associations, however given the small sample size, further analysis in larger populations is necessary to determine the relationships between underlying network activity and brain responses to stimuli.

Finally, a larger (more negative) N1 amplitude was associated with both better verbal and nonverbal measures, suggesting it may be a proxy of overall cognitive functioning. Ethridge et al. 2019 similarly observed that a larger N1 amplitude in response to an auditory habituation task, was associated with better scores on Woodcock-Johnson III Test of Cognitive Abilities Auditory Attention subscale. Authors hypothesized that the increased N1 amplitude within the FXS population may reflect increased hyper-vigilance and in turn improved ability to complete standardized tests.

### Limitations and Clinical Relevance

FXS is a rare genetic disorder, which creates challenges in recruiting large patient samples. In addition, collecting high quality EEG data is challenging in young populations with limited communication abilities and increased behavioral challenges. Despite the small sample size, our findings present a promising future avenue for clinical use. Namely, the VEP P2 component is worthy of continued study as a potential biomarker for FXS itself, as the P2 amplitude was significantly increased in FXS children and often weak or absent in typically developing populations. In addition, this paper contributes to a field of research that is beginning to reveal VEP N1 and P2 amplitudes as a useful neurobiological outcome measure of developmental levels that could be used in clinical trials.

## Data Availability

The datasets used and/or analyzed during the current study are available from the corresponding authors on reasonable request.

## List of Abbreviations

ADHD: Attention Deficit Hyperactivity Disorder
ADOS: Autism Diagnostic Observation Schedule
ANCOVA: Analysis of Covariance
ANOVA: Analysis of Variance
ASD: Autism Spectrum Disorder
EEG: Electroencephalogram
ERP: Event Related Potential
FDR: False Discovery Rate
FXS: Fragile X Syndrome
FMRP: Fragile X Messenger Ribonucleoprotein 1
GABA: Gamma-Aminobutyric Acid
HAPPE+ER: Harvard Automated Processing Pipeline for Electroencephalography plus Event-Related Software
MSEL: Mullen Scales of Early Learning
NVDQ: Nonverbal Developmental Quotient
PLS: Preschool Language Scale
TSC: Tuberous Sclerosis Complex
VEP: Visual Evoked Potential

## FIGURE TITLES AND LEGENDS

**Figure 1: Grand average VEPs and P2 Amplitude across groups**. (A) Grand-averaged VEPs are shown, with 95% confidence intervals. (B) Mean P2 Amplitude across trials across groups. P values shown are in comparison to the FXS group.

**Figure 2: VEP N1 Amplitude and fine motor development**. Correlation between N1 Amplitude and MSEL fine motor raw score for FXS, Age Match, and Cognitive Match groups are shown.

**Figure 3: VEP component amplitudes and receptive language development**. Correlation between N1, P1, and P2 Amplitude and PLS Auditory Comprehension Raw score for FXS, Age Match, and Cognitive Match groups are shown.

## DECLARATIONS

## Acknowledgements

We thank all the families who participated in this study. We also thank Jack Keller, Megan Lauzé, and the Translational Neuroscience Center Human Neurobehavioral Core at Boston Children’s Hospital for their assistance in data collection. We also thank the Infant Screening Study Team for their assistance in data collection of the cognitive-matched controls. We thank Graham Holt for his EEG technical support.

## Author Contributions

CLW contributed to the conception and design of the study. CLW, JF, and MH recruited participants and collected data. ES and CLW performed the data analysis and data visualization, and wrote the draft of the manuscript. All authors contributed to manuscript revision, read, and approved the submitted manuscript.

## Ethics approval and consent to participate

The study was approved by the institutional review board (#P00025493, P000183777).

Consent for publication: N/A

## Author Information

Not applicable

